# A Realist Evaluation of a Fatigue Risk Management Plan (FRMP) Implementation in Obstetrics and Gynecology Residency: Calls for Systemic, Structural and Cultural Reform

**DOI:** 10.1101/2024.11.19.24316016

**Authors:** Aliya Kassam, Benedicta Antepim, Kimberley Thornton, Jason Kumagai, Sarah Glaze

**Author notes:** **Corresponding Author:** Aliya Kassam, Associate Professor, Department of Community Health Sciences & Office of Postgraduate Medical Education, Cumming School of Medicine, University of Calgary.

## Abstract

**Objective:** Accreditation standards in residency education are calling for initiatives to promote the wellness of resident physicians including the implementation of fatigue risk management plans (FRMPs). We sought to conduct a realist evaluation of a FRMP within a five-year Obstetrics and Gynecology (OBGYN) residency training program in labour and delivery units.

**Design:** Mixed method study design informed by realist inquiry.

**Setting:** A single OBGYN residency program comprised of N=32 resident physicians who provided in-house labour and delivery call at four hospitals across a city of over one million people.

**Method:** Realist inquiry askes what works, for whom, in what circumstances, and why? through examining contexts, mechanisms, and outcomes. We collected quantitative and qualitative data from resident physicians sequentially across three time points. Data were thematically analyzed, and configuration on contexts, mechanisms, and outcomes were identified using a realist approach.

**Results:** There was n=19 unique participants (60% response rate). Most of the participants identified as women (93.7%), single (56.2%) and without children (93.7%). Participants mean age was 28.5 years and ranged from junior to senior residents. We found no significant difference between median sleepiness scores across three timepoints (p=0.17) however 20% of residents reported starting and ending shifts with high sleepiness scores signaling impairment that is hazardous to both residents and patients. The n=6 resident interview participants, reported overlooking patient details (forgetting to order tests, and delaying care) as well as personal safety issues such as driving home whilst fatigued despite FRMP implementation.

**Conclusion:** Specific aspects of the OBGYN FRMP such as the nap model and nutrition could help with decreasing perceptions of fatigue related to lack of sleep and lack of available food. While FRMPs might be helpful in de-stigmatizing fatigue in residency, FRMPs are unlikely to decrease levels of resident fatigue because of systemic, structural, and cultural barriers.

## Background and Rationale

The problem of poor well-being in residency has been established.^1-3^ Burnout, depression, and suicidal ideation are prevalent among resident physicians with these issues persisting into practice.^1 4^ Leaders in postgraduate medical education (PGME) have been called upon to create evidence-based initiatives for “wellness” ^5^ including fatigue risk management plans (FRMPs).

There is wide acknowledgement in the literature that the burden of fatigue risk management (FRM) must be shared across all stakeholders in PGME as stated by the Fatigue Risk Management Task Force in Canada and worldwide.^6^ The Royal College of Physicians and Surgeons of Canada (RCPSC) also acknowledges residents and physicians more broadly are susceptible to the effects of fatigue than the rest of the population.^7^ Over the past decade, the National Advisory Committee and Expert Working Group^8^, developed the Fatigue Risk Management (FRM) Toolkit^6^ with guidance from FRM experts and stakeholders in PGME from across Canada.

The FRM toolkit was released in 2018 and is the first national resource for residency programs across Canada. The report provides a “non-prescriptive framework designed to assist clinical learning environments, programs and institutions in developing their own local FRM policies and mitigation strategies”.^6^ Furthermore, the FRM toolkit resource recognizes that the guidelines are not a “one-size-fits-all” and that they should “be adapted to suit local resources and contexts”.^6^ More recently, Canadian PGME accreditation standards^9^ are calling for residency programs to ensure: “The curriculum plan includes FRM, specifically education addressing the risks posed by physician impairment to the practice setting, and the individual and organizational supports available to manage the risk”.^6^

Despite the need for FRMP implementation and national support with respect to resources, research has shown that there is much complexity in conceptualizing fatigue, recognizing fatigue, and measuring fatigue before implementation of a FRMP can even occur. For example, fatigue is a multifaceted phenomenon experienced by residents in different ways and requires management beyond duty hours and adequate amounts of sleep. Furthermore, FRMPs should encompass strategies that are aligned with the resident, the program, and the healthcare system in which they work so that there is buy-in, feasibility and sustainability.^4^

One study showed that fatigue may not be uniformly understood as an occupational threat by residents. For example, they reported that under the current system and structures of PGME where residents form a large part of the healthcare workforce, fatigue is constructed and reinforced by the training environment and culture of medicine ^10^ In residency training, fatigue may be seen as a personal challenge rather than a threat to residents’ own wellness as well as patient safety and as such, the implementation of FRMPs in residency training may not have a substantial impact on resident fatigue because it is understood as “inescapable, manageable, necessary and surmountable and deeply ingrained in the local training culture.” ^10^

Similarly, another study found that health care teams hold contradictory beliefs about how fatigue impacts physician performance and patient care outcomes and called this the “fatigue paradox”.^11^ They sampled physicians, nurses, and senior residents across eight specialties including Obstetrics and Gynecology (OBGYN). They identified that resident physicians may be faced with contradictory messaging of fatigue as an individual resident problem but not for patients or the health care system. Furthermore, the stigma of admitting fatigue may harm patients with few mechanisms to mitigate this harm requires that they must adopt the identity of an infallible physician and maintain that fatigue does not harm patients. They concluded that unless the systemic and structural issues in residency education are addressed the fatigue paradox will continue to be sustained.^11^

As there is a need for more FRM research in residency education and considering FRMPs that go beyond targeting the individual fatigued learner, we sought to conduct a realist evaluation of the FRMP implementation within a five-year OBGYN residency training program.

## Our Focus on Obstetrics and Gynecology Residency Training

The literature commonly shows residents in OBGYN programs in particular encounter unique stressors and have high rates of burnout given the unpredictable nature of obstetrical services.^12 13^ It is estimated that 25%^14 15^ of residents are burnt out and 40% to 75%^14 15^ of practicing obstetricians and gynecologists suffer from work related burnout, making the lifetime risk inescapable.^12-15^ One study found that residents in OBGYN with burnout reported a lack of support by faculty, fellow residents and perceived there is a greater emphasis on service over education. Residents also report a dissatisfaction with hospital benefits and facilities such as call rooms.^14^

Another study found that the number of residents reporting any problem with wellness increased significantly between the first year and second year of training, after which it continued to increase as training progressed. Residents reported programs should provide personal protected time for maintaining wellness and highlighted incongruencies between current system-level solutions and what residents find helpful in maintaining their wellness.^14^ One recent pilot study demonstrated residents, regardless of postgraduate year (PGY)-level or shift, found non-urgent pages to be a significant contributor to on-call fatigue. To manage fatigue risk, a problem board was established to identify and show potentially urgent cases and categorize the cases in a visual way to ensure patient safety.^16^

Given the lack of literature around FRMP implementation in residency education (and specifically in OBGYN residency), as well as the call for FRMPs as part of residency education, we sought to conduct a critical realist evaluation^17^ to understand the effect of FRMP implementation for OBGYN resident physicians at our institution.

## Fatigue Risk Management Plan (FRMP) in Obstetrics and Gynecology Residency Program

The rationale for the FRMP was based on our OBGYN program becoming early adopters of FRM strategies, first by changing weekend call from 24 hours at a time to two 12-hour shifts and second, in 2012, the program implemented a unique ‘nap model’ for junior OBGYN residents. In this model, the on-call resident was relieved of their daytime duties between the hours of 1300-1700 preceding their overnight shift. The residency program also aimed to value wellness by allocating several academic half day teaching sessions for a group wellness activity.

Against the existing backdrop of advocacy for program-level FRM initiatives, our team of authors considered FRM strategies across the levels of self (individual), program and system informed by strategies from holistic wellness, residency education (protected time for academic half-day sessions) and human factors (conceptualization of fatigue, nutrition, and performance) to comprise a comprehensive FRMP for the residents. At the time of the study, the nap model had not yet been evaluated and it was acknowledged that the nap model alone could not be sustainable as the only FRMP. Appendix 1 provides a holistic description of the FRMP in the OBGYN program. We implemented an OBGYN FRMP that targeted all 30 residents across four adult sites with Labour and Delivery Hospital Units in a large Canadian city over the course of one year (2020-2021).

## Reflexivity

Throughout the study and as part of the FRMP implementation the authors each had unique roles in the implementation. The team was led by a medical education and health services scholar working in residency education who assisted with grocery delivery and oversaw the research project. The team also consisted of : a) a research assistant who assisted with study design, ethics approval, data collection and analysis, b) a recent graduate of the OBGYN program and locum who assisted with grocery delivery and was also the FRM champion, c) a human factors specialist who provided human factors expertise and conducted the academic half day sessions on human factors, nutrition, sleep, and fatigue and d) a program director of the OBGYN residency program who facilitated connections with the health care system such as L and D units.

## Methods

This study was approved by the research ethics board at our university.

### Philosophical Approach

The FRMP is a complex intervention meant to be delivered in multiple contexts and settings. As Ellaway et al., indicate “critical realism and its operationalization in the form of realist inquiry can provide much-needed explanatory power” for interventions such as the FRMP. ^18^

In critical realism, the fundamental reason for mixing quantitative and qualitative methods is to promote an understanding of the complexity of the reality.^19^ Furthermore, critical realism assesses the phenomena of existing mechanisms through quantitative data^20 21^ whilst qualitative methods are used in obtaining in-depth explanations of existing mechanisms. ^18, 22 23^ We collected survey, validated psychometric scale data and sleep diary logs as quantitative data and resident perceptions through interviews as qualitative data.

### Study Design

This realist evaluation used a mixed methods design^24^ and involved collecting both quantitative and qualitative data sequentially during multiple points of time. A realist evaluation^17^ (by virtue of realist inquiry) helps to explain “what works, for whom, in what circumstances, and why?” as opposed to “does it work?” As such, the use of realist inquiry seeks to examine contexts, mechanisms, and outcomes. Through realist inquiry, we explored whether a comprehensive FRMP enhances resident wellness because residents may have made decisions in response to the FRMP. Moreover, the reasoning of the residents in response to the resources or opportunities provided by the FRMP may have led to certain outcomes. The mechanisms refer to the underlying social or psychological drivers that led to the reasoning and decision making of residents regarding their own fatigue management, wellness and that of their colleagues (outcomes).^17^

Realist inquiry explains changes (if any) brought about by a complex intervention such as the FRMP under specific conditions (PGY levels, L and D units) within a specific context (across four hospital sites, during COVID-19 pandemic). The residents and the FRMP are embedded in a social reality that influences how FRMPs are implemented and how this impacts resident wellness. The context-mechanism-outcome (CMO) framework is used as the main structure for data analysis in realist inquiry.^18^

Our initial program theory was informed by the previous research conducted around self, program, and system as well as core components of education (academic half day sessions), wellness (specifically holistic wellness as described by the WISHES framework)^25^ and human factors (nutrition, fatigue, and impact measurement of sleepiness). Thus, our initial program theory described how, (in L and D units within the OBS/GYN program), a successful FRMP implementation might be implemented and measured by the Karolinska sleepiness scale (KSS) ^26 27^, Maslach Burnout inventory (MBI)^28^ and resident perceptions through qualitative interviewing. Our initial program theory postulated that within the OBGYN residency training program contexts from PGY 1-PGY5, the FRMP could lead to decreased burnout, decreased sleepiness, overall decreased perceptions of fatigue, and increased perceptions of wellness. Figure 1 summarizes our initial program theory.

**Figure 1.**
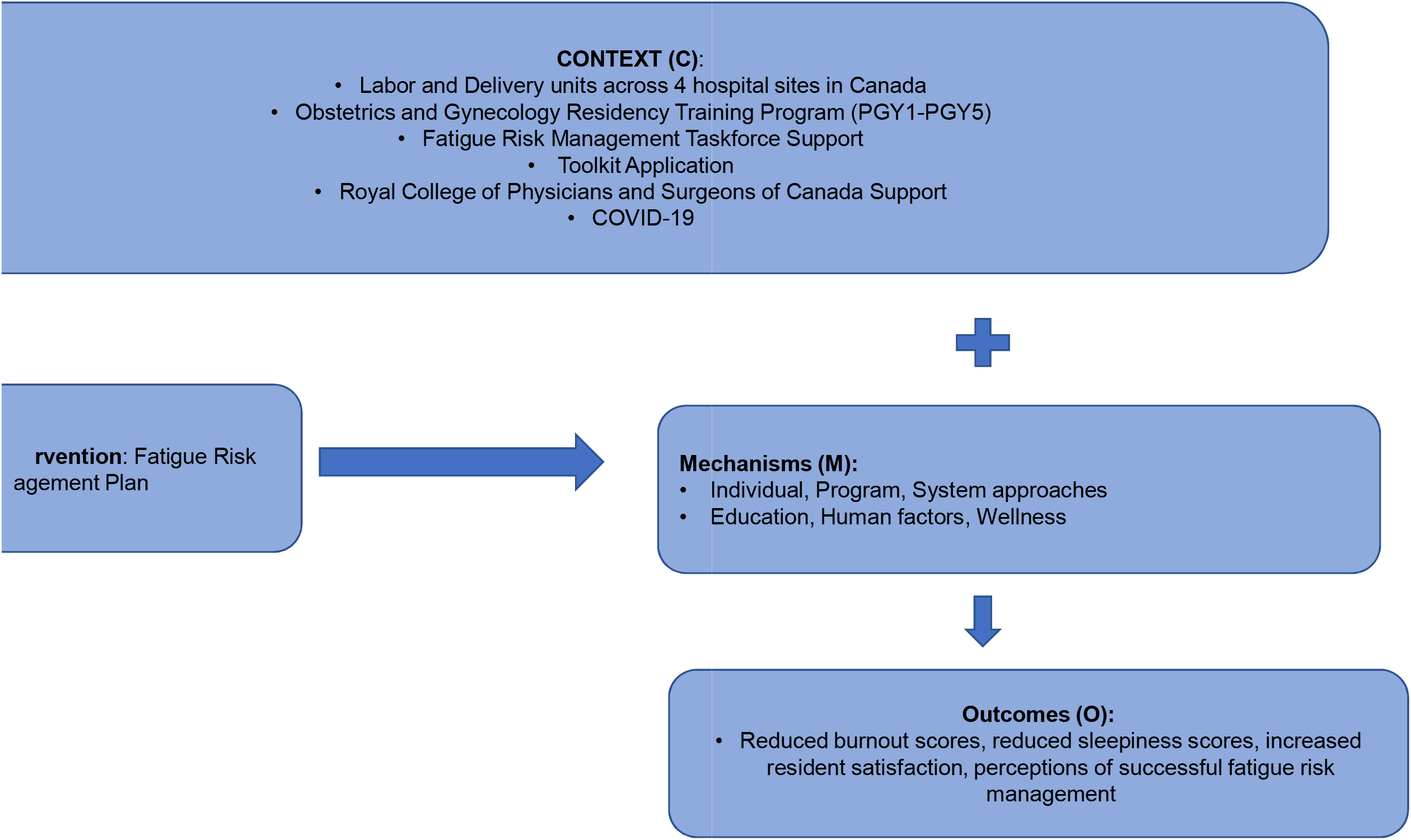
denotes the initial program theory for the Fatigue Risk Management Program (FRMP) implementation.

### Setting

The OBGYN resident physicians provide in-house L and D call at four hospitals across the city. During the study, the breakdown of residents training in the program were (N=32) according to postgraduate year (PGY) was as follows: n=6 PGY1, n=6 PGY2, n=6 PGY3, n=8 PGY4 and n=4 PGY5. Two residents were on leave. The investigators along with systems (PGME and health care system) and national support from the RCPSC recognize the intense demands of the OBGYN residency program, mainly because of the sleep disruption necessary to successfully navigate this specialty.

### Sampling and Recruitment

Authors held an information session about study participation and informed consent during an academic half-day session for the residents. Participants were contacted by the research assistant to participate in the study. Recognizing that the residency program director was also a co-investigator, residents were assured that only the research assistant and principal investigator would know who participated in the study and that participation was completely voluntary. Residents were informed that should they not wish to participate; this would have no bearing on their training and career progression.

### Data Collection

To test our initial program theory, we collected both quantitative and qualitative data.

#### Quantitative data

Residents were asked to complete a pre-mid- and post-test survey (at 0, 6 and 12 months respectively) that assessed and measured burnout using Maslach’s burnout inventory^28^ (which included three sub-scales measuring emotional exhaustion, depersonalization, and personal accomplishment). Residents were also asked about overall wellness, sleepiness, nutrition, and access to food and were asked to complete a sleep diary documenting their perceived sleepiness and sleep quality over a two-week period concurrent with the pre-mid- and post-program mark. A link to the sleep diary was sent by text message to each participant at 7:00 am daily over the two-week period. At the end of the 12-month period of implementation, two research assistants conducted interviews with residents to further explore the outcomes of the FRMP implementation.

#### Qualitative Data

A semi-structured interview guide was developed by the research team and asked about resident experiences of fatigue (both causes and consequences of fatigue), how they managed fatigue as well as their perceptions of the implemented FRMP. Interviews were conducted by telephone or videoconference and ranged between 28 and 61 minutes. Interviews were transcribed verbatim by an independent transcriptionist and all identifying data were removed. Purposive sampling was conducted to ensure we are gathering data from a broad range of diverse perspectives across residents.

### Data Analyses

Survey and sleep diary data were analyzed using SPSS software and we provide descriptive analysis. Interview transcripts were reviewed and coded line by line deductively using the realist inquiry context-mechanism-outcome (CMO) framework using QSR Nvivo software. Regular meetings were held between the research team to ensure discrepancies were discussed.

Throughout the research process from 2020 to 2022, we had regular meetings with the FRM taskforce from the RCPSC and an international subject matter expert. The knowledge dissemination phase has taken place from 2022-present with continuing information sharing meetings to create a community of practice (CoP).^29^ The CoP meetings have been facilitated by the RCPSC FRM teams across Canada and are ongoing at the time of writing this manuscript.

## Results

There were n=19 unique participants over the course of the study (60% response rate). The demographic information of the 16 participants that completed the initial survey showed that participants mostly identified as women (93.7%), single (56.2%) and without children (93.7%). The participant mean age was 28.5 years. Participants ranged from junior to senior residents with n=2 from PGY1, n=5 from PGY2, n=2 from PGY3, n=4 from PGY4 and n= 3 from PGY5.

### Quantitative Data

#### Sleep Diary

Sleep diary data was collected for n=19 participants over the three time points. 54-68% of shifts lasted over 10 hours at every time point. Average length of shift at baseline was 14.0 hours calculated over 129 shifts, at midpoint was 13.0 hours calculated over 85 shifts and at endpoint was 12.0 hours calculated over 50 shifts. Between 12-18% of the shifts lasted 24 hours or longer.

#### Burnout

Across the three timepoints of baseline, mid-point and endpoint respectively (n=14, n=10 and n=7), residents showed median burnout scales indicating emotional exhaustion a few times a month across the three time periods (2.8 vs. 2.8 vs 2.9) whilst residents felt a sense of personal accomplishment between once a week to a few times per week (5.1 vs. 4.5 vs. 4.9). Residents felt a sense of depersonalization between a few times a year or less to once a month or less across the three time points (1.4 vs. 2.2, vs. 1.6).

#### Sleepiness and sleep

We found no significant difference between median scores using the KSS before and after resident shifts on the L and D across three timepoints (p=0.17) during the FRMP implementation, however 20% of residents reported that they were starting and ending shifts with high KSS scores of 8, 9, or 10. Of note, a KSS score of 8 (sleepy, some effort to keep awake) or higher can be hazardous to both the resident and to patients in their care. ^30^

### Qualitative Data

#### Causes, Consequences and Management of Fatigue Findings

There was n=6 residents who participated in the interviews. Supplemental Table 1 describes the causes, consequences, and how residents managed their fatigue. Overall residents reported that the causes of fatigue were related to contextual factors such as being a resident, being on call and the culture of medicine. In terms of mechanisms that were fatigue-related, residents reported decreased emotional regulation impacting their mental, physical, and intellectual wellness. These led to outcomes that impacted patient safety such as overlooking details, forgetting to order tests, and delaying care as well as personal safety issues such as driving home whilst fatigued. In contrast residents reported positive outcomes to their social wellness because of mechanisms such as residency program cohesion:

> *“I totally love my resident cohort. And I feel I have made five soul mate friendships in residency. So, I feel that’s been really great. And the rest of the program has been so supportive. And the staff and my seniors are all just such lovely people that I feel socially are very well connected.”* – Participant 02

There was also a sense of appreciation that the program was being proactive about FRM which also acted as a mechanism:

> *“I think it’s very good that the program is working on understanding the residents’ fatigue and making it better and trying to work on patient safety because I think, in talking to other residents in other programs this isn’t something that gets talked about and so I think that this is super-important.”* – Participant 03

Regarding occupational wellness, there were mixed responses. On the one hand increased covid-19 restrictions in the context of the pandemic allowed time to take care of oneself leading to a positive outcome:

> *“So for example, prior to COVID we would never take any time off, even if we were ill. So, everyone has examples of when they were just so sick, but still came to work. I remember I started getting a lot of muscle aches and pains, and neck pains, especially towards the end of residency, and it was really difficult to find time to recoup around that, because you still had to go to work and perform.” -* Participant 04

On the other hand, residents were left feeling more exhausted because of the mechanism of increased workload, clinical complications, and uncertainty of acquiring skills for competence:

> *“There’s some pieces of COVID that are just, you feel constantly, clinically exhausted, because we’re dealing with really sick people and really bad complications because of COVID. And that’s really hard on people. That’s really hard on residents. I think it’s been hard because there’s also uncertainty about acquisition of skill, especially in terms of our competencies.” -* Participant 05

Management of fatigue by residents as outcomes were reported as being individual level strategies such as caffeine, exercise, therapy, making notes and lists, and antidepressants.

#### FRMP Evaluation Findings

From the quantitative data presented above, we found that the interventions of the FRMP had little tangible impact on fatigue, in terms of reported sleepiness, actual time slept, or burnout. The qualitative data and realist evaluation makes it clear that the interventions were successful in improving the perception of fatigue among some residents but not the fatigue itself. Table 1 provides a summary of the FRMP context-mechanism-outcome configurations (CMOs).

**Table 1.** Context-Mechanism-Outcome (CMO) Configurations of the Fatigue Risk Management Plan (FRMP)

| Context | Mechanism | Outcome |
| --- | --- | --- |
| **Junior residents** | R1-R3 experience → physical fatigue from long hours/call shifts, emotional fatigue. | Fatigue was mitigated by interventions that addressed key sources of fatigue – for example nap model for R2s, fridge and snacks for call rooms. |
| **Senior residents** | R4 and R5 experience → mental fatigue, intellectual fatigue, better balance, and control. | FRMP interventions had little impact as none directly addressed sources of fatigue at systemic, structural or cultural levels. |
| **Residency program** | Overwhelming and overprioritized | Individual level strategies to reduce impact of fatigue |
| **COVID-19 – Disruption of schedules** | (+) Less time at work<br>(-) Educational milestones<br>(-) Disrupted emotional state and coping | (+) More time to engage in self-care.<br>(-) Loss of social supports |

(+) Denotes a positive aspect in the mechanism or outcome.
(-) Denotes a negative aspect in the mechanism or outcome.

Based on the recurring patterns that are evident throughout the CMO configurations, we can theorize that for the FRMP interventions had a positive impact (and may work in other contexts), with the nap model and nutrition initiatives seeming to have had the most impact. Overall, the FRMP was an improvement to the resident program experience and was perceived to have positive impacts on perceptions of fatigue, so long as they aligned with already existing individual level mitigation strategies and sources of fatigue. Supplemental Table 2 highlights perceptions about the FRMP initiatives.

## Discussion

Our study had three main findings. First, despite best efforts of implementation of a FRMP (as per accreditation requirements) for OBGYN residents, there are factors that impede its effectiveness that are inherent to residency such as, long hours, call shifts, a high stress environment, and the culture of medicine. FRMPs at the program level to address fatigue are only a band-aid solution. Second, it remains clear that resident fatigue can have significant impacts on patient and personal safety, this was reported by the residents in this study as well as corroborated by many other studies on this population and others.^10 11 16 31^ Last, we found that FRMP interventions that are able to address the systemic, structural and cultural concerns that cause these issues must be aligned with the program level FRMP.

We cannot understate the importance of recognition and awareness by the OBGYN program in implementing the FRMP and the consensus is that while FRMPs might be helpful in de-stigmatizing fatigue in residency, such interventions are unlikely to decrease levels of resident fatigue because of the systemic and structural barriers as well as the culture of medicine. Given that programs taking steps to facilitate and provide space and resources for wellness and coping strategies helps improve the perception of program support and improves program culture, our findings align with previous studies that while FRMPs might be helpful in certain aspects of fatigue, they may not be holistically helpful. ^10 16 31^ Additionally, as residents move through residency, they might become more resilient in coping with fatigue which then impedes innovation and the call for systemic, structural and cultural change in medicine. So where to go from here?

Residency education and the health care system has to grapple with the issue of liability when it comes to fatigue. Our results showed the increased risk of impairment in fatigued residents which could lead to patient safety and/or personal safety issues. In these cases, we must then ask how can adverse events to both patient and resident be avoided if fatigue is driven by the very nature and key features of how we conduct residency education. Furthermore, who is liable when we inevitably encounter adverse outcomes as a result of residency training?

In terms of lessons learned from our realist evaluation, we have now refined our initial program theory (see Figure 2) to encompass systemic and structural barriers as well as culture that could impede program level FRMP implementation. We also would like to re-iterate^31^ that FRM is a multi-facetted issue with individual differences and ultimately residents rely on their own FRM strategies to help them through residency.

**Figure 2.**
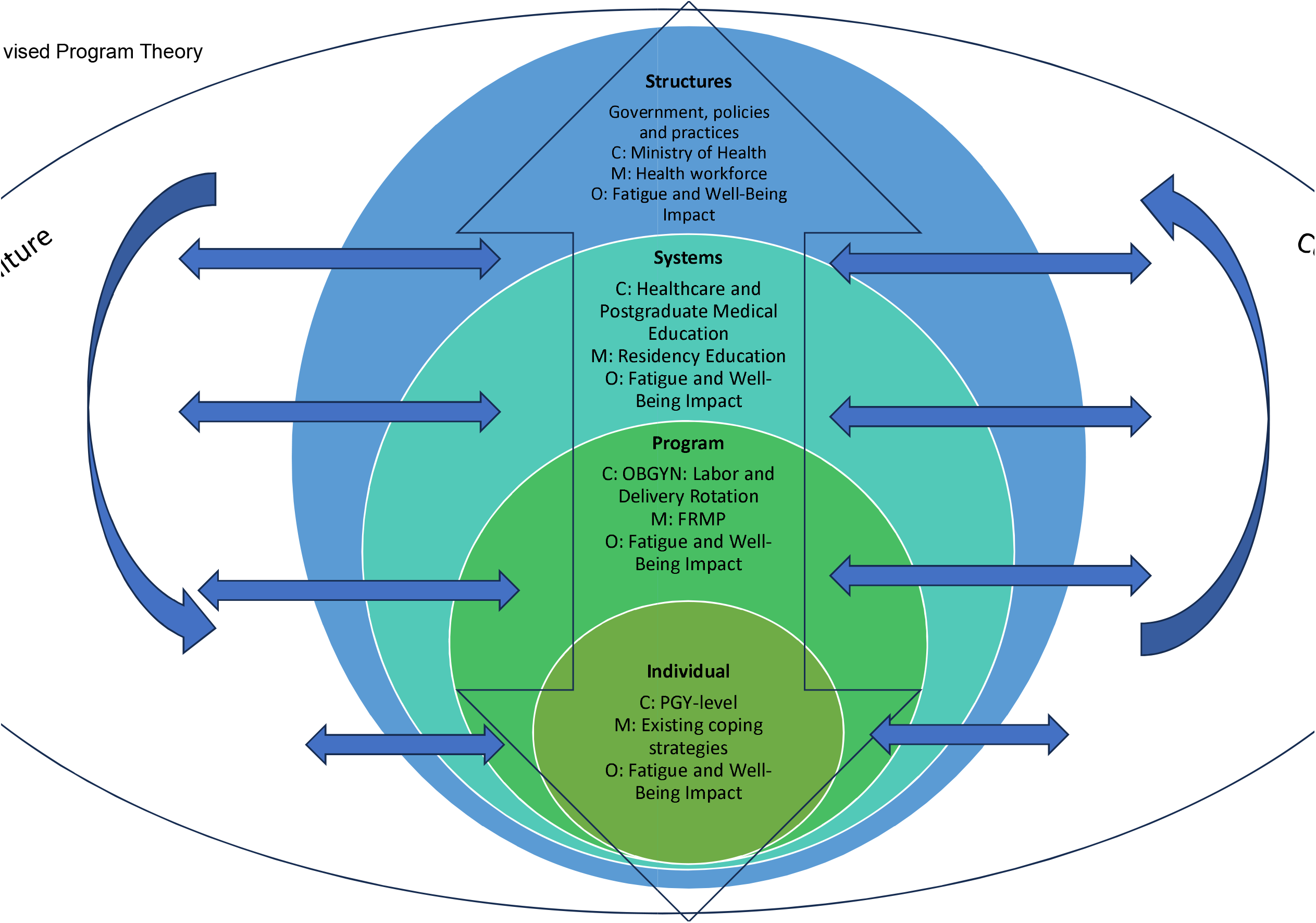
denotes the refined program theory from the initial program theory to encompass systemic and structural barriers as well as culture that could impede program level FRMP implementation.

In terms of best practices from the FRMP implementation, we suggest normalizing the nap model (so that residents are not reluctant to use it) and allow protected time for self-care, list clear tasks for the FRM officer (such as regular check-ins) and provide call room nutrition by way of having a fridge and snacks available on a regular basis. Future research should look at the intersections of fatigue and competence assessment while considering contextual factors such as relational autonomy, emotional tone, socio-cultural aspects as well as technology.^32^ Future FRMP implementation should also apply complexity theory and implement FRMPs considering complex adaptive systems.^33^ This would help in considering the bigger picture in which the FRMP interventions are implemented.

A major strength of this study is that it is the first to use a realist evaluation for implementation of a FRMP. Despite this, there were several limitations to this study. First in collecting several indices of data, there may have been research-induced fatigue which could have increased the fatigue experienced by residents. In consulting with an international sleep subject matter expert, we had to increase the data collection instruments and make changes to the sleep diary questions across the data collection period which may have impacted the results and induced further fatigue in residents. Second, we had a small sample size with loss to follow-up across the time points which could compromise the generalizability of our results. Only 19 of the 32 residents participated, and even with this sample, we had significant attrition from baseline to endpoint. Another key limitation is that we used self-reported sleep and work diaries, which introduces recall and response bias which could be further confounded by being fatigued. Last, COVID-19 changed the context of this study and did not represent what a usual environment looked like for residents thereby also impacting levels of fatigue.

CanERA General Standards of Accreditation for Residency Programs require *“*effective central policies and processes are in place addressing residents’ physical, psychological, and professional safety”, including FRMPs ^9^. Studies like ours demonstrate the challenges of culture change but also provides examples of achievable and meaningful interventions in FRMPs that residency programs can aim to accomplish. Improvements to existing FRMPs, include regular sessions to help de-stigmatize talking about fatigue, increased outreach to residents by the FRMP officer, in-house seminars by staff who understand systemic, structural and cultural barriers and sustained food and nutrition initiatives.

Our study showed findings that refuted our initial program theory that the FRMP would decrease burnout and fatigue. However, we also found aspects of the FRMP such as the nap model and nutrition could help with decreasing perceptions of fatigue related to lack of sleep and lack of available food. Future FRMPs should examine the role of the FRM officer and explore contextual features to specific training environments within residency programs such as L and D hospital units as part of OBGYN residency and how various mechanisms moderate outcomes using realist evaluation.

In conclusion, resident physician FRMPs cannot be implemented in isolation without consideration of the broader systemic, structural and cultural influences which modulate various contexts, mechanisms and resulting outcomes.

## Supporting information

Supplemental Tables with findings

## Data Availability

All data produced in the present study are unavailable since data sharing was not approved by the research ethics board.

## Acknowledgements

We would like to thank Dr. Drew Dawson for his fatigue risk management expertise and advice in measuring sleep and sleepiness. We would like to thank the Royal College of Physicians and Surgeons of Canada Fatigue Risk Management Taskforce for funding this research.

