## Supplemental Tables with findings for "A Realist Evaluation of a Fatigue Risk Management Plan (FRMP) Implementation in Obstetrics and Gynecology Residency: Calls for Systemic, Structural and Cultural Reform"

Supplementary Table 1: Causes, Consequences and Management of Fatigue

| **Causes of Fatigue** | **Supporting Quotes** |
| --- | --- |
| Residency | “My physical wellbeing is definitely worse than it was when I entered residency. I feel there is not as much time to go to the mountains, go hiking, not as much time for just working out….” Participant 02  “….you just like abandon your hobbies. Or just because you spend so much physical time at work, and then you had so much limited time at home, that you needed to recoup, and therefore like that time had to come out of something, which as we know, time is a finite resource, and there just wasn’t enough of it to go around. Participant 04  “So, things look a little bit different I would say as a junior [resident], you’re definitely really more emotionally fatigued because it's the first time you’re dealing with a lot of stuff and there’s a lot of learning that happens really fast and you’re trying to keep up and perform.”  Participant 05  “….you know, actually being on L&D and obstetrics for 12, 13 hours is very physically demanding and so you’re often, like not just emotionally and mentally, but like very physically drained too, at the end of the day.” - Participant 06 |
| Being on call | “….one of my colleagues when in the throws of doing a lot of call in the week, fell asleep on call and then didn’t answer their pages so then there’s nurses paging about things that like are going unaddressed; and so, if those things are urgent then that’s bad.” - Participant 03  “I would say definitely the physical hours, because you easily work 80-hour weeks, and that doesn’t leave a lot of room for kind of replenishing yourself.” Participant 04 |
| Culture | “And then there's been some periods where you're oh, is medicine at all even the right place for me? Or are there yeah, there's been some bad outcomes that I've had that have just kind of lingered and been quite traumatic for everybody that's been involved, including myself and so those have definitely been some low points.” Participant 02  “And then I think also one of the things about residency, that there’s a lot that is hard to argue around, is that everyone says you’re a trainee, and so it really is about getting all – as many hours and experiences as you can in, before you’re out to practice. And I found it really difficult to challenge any notion of, you know, getting more time off to rest, or getting more time off to recoup, because you’re constantly thinking – you’re like, no, this is my training, I have to put in as many hours as I can, this is an expectation of me.” Participant 04 |
| **Consequences of Fatigue** | |
| Impact on patient care | “I think that there's probably lots of times when you're rounding on your post call, or really tired, and you probably aren't as thorough in evaluating a patient as you'd like to be. I think also I definitely I'm right now I'm in a rotation where you do home call, and you kind of get woken up in the middle of the night. But you're more likely to be asleep, so you're kind of groggy and I think there's been a time when I had a consult from emerge that I probably didn't, I guess pay close enough attention to. And I wasn't really pleased with how I handled the patient in the end. Just I think I just because I was tired and trying to get some sleep. I probably didn't pay close enough attention and didn't put a plan in place that I was very happy with. Both overnight and then in the morning when I saw the patient. Probably the most specific example I can give I think is mostly just I feel missing things, missing lab results or missing a new symptom or something that when you're tired, you might just kind of not hear or not ask about.” – Participant 01  “I think things get missed. So, whether it's lab results, a patient who might need a blood transfusion and you're waiting for their haemoglobin to come back. Or patients who have preeclampsia and you're monitoring their labs, and maybe debating if you need to start a different medication. Even just I find the main point of safety that is that routinely comes up is at the point of handover. Or it's those little things that you don't have somebody haemorrhaging in front of you. But it's those little things particularly on the unit because they're kind of out of sight and tucked away that get missed. And so, someone might have a very high blood pressure on postpartum and no one's alerting you or there might be different kind of critical results that aren't immediately related to you.”  – Participant 02  “I can think of one example where a patient – that happened, and a patient’s C-section got delayed until the morning, and on that baby, the tracing had been fine before that happened, and she was a low-risk patient, so they took her off the monitor, and they let the patient rest. And end of the next morning when they did the C-section, the baby had in fact been in distress. And I can’t remember if the baby passed, or had really poor neurological outcome, yeah. But I mean obviously specifically in that example, who really knows what the issues were, but I do think part of it was no one wants to do something in the middle of the night at three am, yeah, if they think that they can help it. And I think it if had been obvious that the baby had been distressed, obviously people would have acted on it. But when people perceive that there is not as much urgency, I think then – but when the manifestations of people’s fatigue start to become noticeable on patient care.” – Participant 04 |
| Impact on personal Safety | “So overall I know nightshifts going to cut years off our lives, we are definitely not as healthy when we work nights as we are when we don’t work nights for sure. I know there’s lots of residents who’ve gotten in car accidents on the way home from call, I haven’t had that happen to me luckily but I'm sure it could. And your physical safety when you’re so tired, I don’t do any high-risk sports when I'm post call the stuff like that I would normally do I don’t do any of that when I could, your reaction times a lot slower for sure. And I think just physically you’re not as fast, you don’t respond as well so definitely in that.” Participant 05  “And I think we had three residents in the last year that were in car accidents on their way home from work. So I feel it would - I know, of residents in general surgery two in the last year that were in a car accident on their way home. So it's actually really common, that people are getting into accidents. And I myself has fallen asleep multiple times driving home.” Participant 02  “I've definitely fallen asleep at a red light before and then woken up when someone beeps somewhere. We all sort of joke about that which is very dangerous but its better now than it was before, I'm less tired after a night than I was when I was younger, in the immediate sort of aftermath. But yeah definitely, I mean we’re up for 24 hours there’s – no matter how much intervention I think we’re always going to be tired when we finish 24-hour shifts.” - Participant 05  “So, I think fatigue contributes to all that and kind of chipping away at maybe your resiliency. But, I mean, like obviously physical and personal safety, like driving home from call shifts and stuff, is probably the most dangerous thing I do in my life sometimes, it feels like, but – yeah. I don’t know what else in terms of safety. Maybe, like personal safety, other than, like emotional, physical wellbeing. I think probably – I feel like people – a feeling like it’s taking away years off your life, you recognise that in the moment, but – I mean I don’t ever feel like I’m in danger, I guess, from fatigue, other than maybe feeling very tired on drives home.” – Participant 06  “Well, I think like we drive home from call all the time and so we’re very tired, and then driving while doing that I think is bad. I find like there are things that we can do like we can get taxis home from the hospital, just kind of like a hard system to use because you can’t taxi to the hospital and home, you have to like taxi home and then back to so your car get trapped at the hospital; so like safety-wise that way.” Participant 03 |
| **Managing Fatigue** | |
| Snacks and hydration | “Yeah, I mean, like I think certainly the snacks, to be honest with you. Having any form of just sustenance or nutrition, I realise I think it makes you much less fatigued the next morning. Yeah, I think on nights where you don’t sleep, how drained you are, and I’m sure it’s impacted by even having a small snack or a bite to eat or something. I think it makes a big difference. And hydrating too. I don’t hydrate enough. So, like having a sparkling water or something there was great, because we often forget to also drink any water during the day. And then I think that drains you as well.” – Participant 06 |
| Making notes and lists | “I start using a lot of lists and more binders and highlighters to make sure that I'm trying not to miss the things that either I want to hand over to the oncoming team, or in terms of some of those things that are a little bit easier to forget like this referral that I forgot to put in. Yeah, so I revert to a lot of write things down, and checklists.”  Participant 02 |
| Caffeine | “I guess I mean, we all drink, well, most of us drink coffee or tea, so caffeine to try and stay alert and minimise patient safety issues.” - Participant 01  “I just drink a lot of coffee. Like a lot of coffee. And then I sleep pretty much all my post call days, some people are very productive, but I'm somebody who comes home and sleeps the entire day.” – Participant 02 |
| Setting realistic expectations | “But I think just like being aware of fatigue and where it’s coming from and like things that you can do to mitigate it or at least just being, again, like kind with yourself and knowing that this is fatigue, I need to actually put effort and mindfulness into dealing with it and not just hoping it will go away on its own. I think like that will carry forward into like being a staff and those type of things. And also just like remembering how hard residency is moving forward into like one day when I am a staff just like having kindness and forgiveness and like patience with residents who will generally be up many more hours a week than we are as staff.” Participant 03 |
| Sports and exercise | “Well, I definitely use exercise quite a bit I and I find that that's a big factor for me. If I don't exercise, I do notice a change in my mental health and my emotional state as well. So that's a big one for me in terms of wellbeing.” – Participant 01 |
| Therapy and antidepressants | “So, I had to get a therapist, I got myself on antidepressants, I had to talk to my colleagues, a lot of who experienced the exact same thing.” Participant 04 |

Supplementary Table 2: Perceptions about the Fatigue Risk Management Plan (FRMP) after Implementation

| FRMP Intervention | Supporting Quote |
| --- | --- |
| Nap Model for 2nd Year Residents | *I think it was helpful. I mean it was really helpful in a sense that at least I got bit of a break. Like I definitely preferred it to not having it, yeah. But I think it was really interesting, because I think the unintended consequences of it was like a lot of guilt, and almost shame, for reminding people about the Nap call, guilt because, you know, you were leaving your colleagues when it seemed like it was really busy, and you felt like you couldn’t help them out.* – Participant 04  *When we had that nap call system I always slept before call and that really helped in second year. Now if I can do it I will but usually as a senior I don’t have the opportunity to take any time off before I go to call. Because I find for me anyway really the call is what gets me, long hours, early mornings I don’t really, I don’t get fatigued as much by that, that I can handle.* Participant 05 |
| Nutrition Initiative | *I've loved the programme for the snacks and the fridge that was provided. And I think that's been actually a really great thing. In terms of keeping us energised, and also keeping us eating something reasonably healthy. And the fridge is great, because it encourages people to bring their own food too so you can eat a bit better.* Participant 01  *Having healthy granola bars and like those types of things accessible is really helpful because like we’re so busy we don’t always have time to go and buy these types of snacks for ourselves. So just being able to have access to them at all times of day is helpful because there’s a lot of times where we’re so busy that the first time you get to eat in a day is 2 am and so then it’s too late to get anything, so having that available is good.* - Participant 03  *But one thing I did appreciate was the healthier snacks in the call rooms, because I found sometimes those were the only things that I would eat all day.* - Participant 04  *I can't speak highly enough about the fridge and the food that has greatly impacted things for me.* Participant 06 |
| Nutrition and Sleep Hygiene Seminars | *The coaching sessions I enjoyed in the moment, but I probably haven't retained as much of that information as they would to or should have. And so, I probably have not been able to implement those, I think maybe if they've been maybe broken down into several sessions, instead of just two at the beginning, it might have been easier to kind of be reminded or to come back to an implement. But I remember enjoying them, I just don't remember the content as well as I would like.* Participant 01  *I thought the sleep talks were actually helpful. In some ways the talk wasn't as applicable to us because we talked a lot about structured napping, but we just can't when we're doing 24 hours, it's just not possible. But I did take away some tips about trying to manage myself post call and when I'm fatigued otherwise, so that was helpful. I did also take away some stuff from the nutrition talk. So, I've changed the times that I eat carbs on call and sugar. So that kind of stuff was helpful. Having snacks always is nice, because at least I knew coming into a call shift. I didn't have stuff with me, then I would have stuff there.* Participant 05 |
| Fatigue Risk Management Officer | *The FRM Officer is just somebody who reaches out on their own sometimes. For example, I had a very bad case outcome. And they just texted me and [they have] just been a good lifeline and support at different times. But I have not necessarily been using them in their formal role [as FRM Officer]. I miss email sometimes*. Participant 02  *I didn't actually know that they were assigned as an officer.* Participant 05 |
